# Benchmarking Structural Variant Detection in Whole-Exome Sequencing of 67 Multiple Myeloma Cell Lines

**DOI:** 10.64898/2026.11.06.26367303

**Authors:** Lokesh Pugalenthi, Rahul Nanduri, Raymond Hong, Maya Ylagan, Rohit Prasad, Dhivya Arasappan, Jeanne Kowalski-Muegge

## Abstract

**Background:** Although the majority of structural variant (SV) workflows have been derived based on whole genome sequencing (WGS) data, they continue to be applied to whole exome sequence (WES) data for which many studies have evaluated their performance in cancer. An equally important consideration that has not been well studied is the impact of cancer context specific features among WGS-derived SV callers applied to WES data. Like personalized medicine, not all SV callers are alike and optimal for use with all cancer types and sequence data types. In addition to sequencing related data features, cancer types have embedded features withing the sequencing data that should be considered. Herein, we performed an analysis of SV heterogeneity among WGS-derived SV callers to WES data in multiple myeloma (MM), a cancer known for its genetically heterogeneous nature. Using WES data derived from a panel of 67 Multiple Myeloma (MM) cell lines, we evaluated seven WGS-derived SV detection tools (Delly, SvABA, BreakDancer, Smoove, LUMPY and Manta) for their ability to identify cytogenetically known genetic abnormalities and to characterize overall differences in the number, length, type, and runtime of SVs called among them.

**Results:** Among the seven tools, Smoove and Manta both had the shortest runtime, processing files in less than 2 minutes, in contrast to tools that required more than two hours to run such as SvABA and Pindel. Smoove was the only SV caller to identify a range of SV types, while SvABA was the most restrictive, calling only breakends. All tools differed in the length of SVs called, with SvABA’s unfiltered output including the shortest length of less than 10 base pairs (bps), and Delly and BreakDancer’s output including the longest, with SVs of less than 1,000 bps. At the chromosome level, SvABA’s unfiltered output and Delly’s output included 78% and 27.5% of known SVs, respectively, with the remaining tools (including SvABA’s filtered output) achieved recall rates of less of less than 10%. By comparison, at the genomics coordinate level, no tool identified more than 20% of known SVs.

**Conclusions:** As all evaluated SV detection tools achieved poor recall rates on this comprehensive catalog of MM cell lines, this study implies SVs detected by WGS-based SV callers on WES data should be confirmed experimentally before being declared legitimate. Heterogeneity in these tools’ output lends support to their combined use in practice. Considering that many clinical grade sequencing panels continue to expand their repertoire of rearrangements with therapeutic implications, our results prompt the potential need for cancer type-matched workflows to evaluate the calling of such rearrangements.

## Background

In contrast to Whole Genome Sequencing (WGS) data, the application of existing structural variant (SV) workflows to Whole Exome Sequencing (WES) data has not been rigorously benchmarked^1^. This gap in benchmarking is particularly concerning with the increased use of WES data to perform clinical grade sequencing for molecular profiling of a cancer patient’s tumor to inform on treatment.

Current literature on WES-derived SV calling using the specific callers Delly^2^, SvABA^3^, BreakDancer^4^, and Smoove^5^, illustrates differences in how often each tool is benchmarked. On the one hand, the application of Delly and SvABA has resulted in a wide range of results in at least 17 studies^1,6-22^; while some studies claim that Delly and SvABA have a low recall of clinically validated SVs, others claim the opposite. On the other hand, there is a scarcity of studies in which BreakDancer and Smoove are applied to WES data; prior benchmarking of their performance was predominantly evaluated on WGS data^23-25^. These differences motivate the need for more extensive analyses comparing several SV callers and their independent benchmarking on WES.

Herein, we evaluated seven SV detection tools on WES data derived from a panel of 67 Multiple Myeloma (MM) cell lines. Cell lines in general are a valuable model to use for such evaluation considering their known cytogenetic-based SVs independent of next-generation sequencing (NGS) data. MM in particular is known to have SV biomarkers of prognosis and treatment response^24^. Thus, there exists a critical need to benchmark these tools and identify reliable SVs in this cancer. Using WES data from 67 MM cell lines, we evaluated seven SV detection tools for their ability to identify known cytogenetic abnormalities: Delly, SvABA, BreakDancer, Smoove, LUMPY, Manta and Pindel.

## Methods

For a summary, see **Fig. 1**.

### MM Cell Lines Description

Raw FASTQ files from WES were obtained on 67 MM cell lines from Dr. Jonathan Keats^26^. Mean read depth at each position was calculated as approximately 13, determined by executing samtools^27^ depth across all BAM files. This low coverage level may reduce the likelihood of capturing multiple independent split or discordant reads required by callers (BreakDancer, Smoove, and LUMPY) to pass high-confidence detection thresholds

### whole-exome sequencing data processing

Bwa^28^ aligner was used to align samples to the GrCh38 reference genome. Indexing and sorting bam files were conducted using samtools^27^ and in-house scripts developed for parallel processing of files using the Texas Advanced Computational Center (TACC) computing nodes. Read group tags were removed and replaced with a ‘missing RG’ tag using GATK^29^ v3.6 to indicate that the BAM files must have no read groups. The modified BAM file was reindexed using BreakDancer, Smoove, and LUMPY.

### Structural variant (SV) pipelines

For each pipeline, we summarize BAM file processing and SV calling method. All pipelines except BreakDancer output standardized variant call format (VCF) files, facilitating comparison of breakend records across pipelines. BreakDancer’s output resembled a VCF file to the extent that comparison with other pipelines was possible.

#### SvABA^3^ (v1.1.0)x

SvABA was directly applied to the unmodified BAM files. SvABA uses local assembly in a way that combines classes of variant-supporting reads into a single framework. For filtering, SvABA performs a series of log-likelihood ratios that are used to threshold variants. Variants with a log-likelihood of at least six were retained for analyses.

#### Delly^2^ (v0.7.6)

Delly was directly applied to the unmodified BAM files. Variant calls of low quality or with base-pair length exceeding one million were removed. For each BAM file, Delly computes the default read-pair orientation, and the paired-end insert size distribution to define non-translocation SVs (e.g., deletions, insertions, and duplications). Delly defines translocations by building a graph of the paired ends. SV filtering is done using a k-mer-based technique, of which the default setting was used.

#### BreakDancer^4^ (v1.4.5)

A BreakDancerMax Perl script was run on modified (i.e., no RG tags) BAM files to produce a configuration file containing key information (e.g., platform, read length, mean and standard deviations) from which nine cell lines (KMS20, JJN3, Delta57, KMS11, Amo1, RPMI8226, MM1S, NCIJ929, MOLP2) were defined to have coefficients of variation ranging from 46.9 to 90.3, and thus, greater than the analysis capability of BreakDancer. Using this configuration file, a .ctx file was derived, a format that resembles a standard VCF file to the extent that it was possible to run the gene annotation tool. Mapped read pairs were classified into six SV types derived from regions with at least two above average anomalous read pairs (ARPs).

#### LUMPY^5^ (v0.2.13)

Discordant paid-end alignments were extracted by calling *samtools view* on the modified BAM files. Split-read alignments were extracted through the provided bash script *extractSplitReads_BwaMem*.*sh* and the *samtools view* command. Both alignments were sorted using samtools, and lumpyexpress was called on the splitters and discordant bam files to produce a VCF. Like SvABA, LUMPY’s SV calling method integrates multiple SV signals. No subsequent filtering done; the creators of LUMPY devised the tool Smoove to provide more filtered output.

#### Smoove^5^ (v0.2.5)

Two Smoove commands were called on the modified BAM files to produce VCF files. The ‘call’ command identified SVs. The ‘genotyped’ command provided statistics regarding alternate alleles. Smoove’s method is akin to LUMPY but with extra internal read-filtering that included discarding inter-chromosomal reads with greater than three matches and reads from regions where the depths of split or discordant reads are greater than 1,000.

#### Manta^30^ (v1.6.0)

A configuration python script was applied to unmodified BAM files. Among the SV pipelines evaluated, Manta was the only tool whose approach was predominantly graphical. All paired and split-read evidence were consolidated into a quality score under a germline model and complemented by filtration metrics to improve call precision. These filtration methods include the following: a minimum alternate allele frequency of 0.3; removing reads marked as secondary, supplemental or PCR duplicates; removing edges in graph with insufficient evidence.

#### Pindel^31^ (v0.2.5b8)

Pindel was directly called on the BAM files. To convert the Pindel raw output into a standardized VCF output file, we used the package pindel2vcf.

### Annotation

SnpEff^32^ (version 5.0) was applied to annotate each VCF file with GrCh38 build.

### Analysis

#### Recall metrics

A measure of recall within each pipeline was computed using curated, known inter-chromosomal SVs in MM cell lines that were computed at both the chromosome and genomic coordinate level. Recall at the chromosomal level is determined by whether for each curated, known SV, a tool identified an inter-chromosomal SV with the same starting and ending chromosomes. Recall at the coordinate level takes this definition a step further by requiring the starting and ending coordinate to be within an apriori specified margin of error. A measure of recall among pipelines was derived by defining the number of tools shared within 100 base pairs.

#### Visualizations

The R BioCircos^33^ package (v0.3.4) was utilized to generate Circos plots showing inter-chromosomal translocations for all characterized SVs; the expression option of UpSetR^34^ was used to visualize the overlap of genes.

#### Clustering

K-means clustering was implemented using ScikitLearn and the optimal number of clusters was determined based on an elbow plot.

## Results

Unless stated otherwise, results and discussion concerning SvABA refers to its unfiltered output, which achieved a higher recall rate than its filtered rate (see subsection Comparison of tool recall performance).

### Comparison of tool output

We compared each tool in terms of the number, type and length of SVs identified, and their runtime. With regards to the number of SVs identified across among the cell lines, all tools except one, Smoove, identified hundreds of thousands of SVs; Smoove detected thousands of SVs (**Fig. 2a**). In terms of SV length, SvABA identified the shortest length SVs of less than 10 bps, in comparison to Pindel and Manta which identified SVs with less than 100 bps, while Delly and BreakDancer identified the longest length SVs of less than a 1,000 bps (**Fig. 2b**). Smoove identified SVs ranging in length from 10 to 10,000bps. Altogether, these results demonstrate the heterogeneity in SV length among tools, lending support for their combined use.

In addition to SV length, we also observed heterogeneity among tools in terms of SV types detected. (**Fig. 2c**) shows this for the tools SvABA, Smoove, Delly and BreakDancer. Breakends were detected by SvABA, Smoove and Delly (i.e., not BreakDancer). Deletions and Inversions were detected by Smoove, Delly and BreakDancer; note that SvABA only detected Breakends. Duplications were only detected by two tools (Delly and Smoove). BreakDancer detected additional SV types: inter-chromosomal (CTX) and intra-chromosomal translocations (ITX).

In terms of the distribution of SVs detected, both Delly and BreakDancer were similar. While they both detected several types of SVs, they report a greater proportion of deletions as compared to other tools. SvABA was the most restrictive, identifying only breakends. In contrast, Smoove resulted in a more uniform distribution of SV types detected. Thus, the heterogeneity in SV types detected among these tools lends further support for their collective use.

Except for Manta and Smoove, all tools showed a similar, significant (p ≤ 0.05) correlation between read counts and runtime (**Fig. 3**). Smoove was the only tool that lacked such a correlation (p=0.21), though Smoove and Manta had the shortest runtime, processing all cell lines in less than 2 minutes, followed by Delly and BreakDancer tools that processed files in less than 20 minutes. SvABA and Pindel tools required the longest runtime, requiring more than 2 hours to process files (**Fig. 3a-b**). When clustering the tools’ runtime across all cell lines (**Supp Fig. 1a)**, 5 was judged to be the optimal # of clusters (through the elbow method and the technique of silhouette analysis). The first cluster would consist of the tools Manta and Smoove as both have relatively short runtimes. The other 4 clusters would each consist of one of the 4 remaining tools (SvABA, Pindel, Delly, BreakDancer).

When clustering each cell line’s runtime across all tools, 4 clusters of cell lines were judged to be the optimal # of clusters (**Supp Fig. 1b**). This means that there are 4 groups of cell lines with distinct runtimes. Each cluster’s cell lines are indicated in **Supp Table 1**. Of particular interest, one of the clusters consisted solely of the cell-line Karpas417; thus, the cell line Karpas417 had the most unusual runtime relative to the other MM cell lines

### Comparison of tool recall performance

At the chromosomal level, SvABA’s unfiltered output achieved a recall rate of 78% (i.e., identified 78% of known SVs). Delly achieved a recall rate of 27.5%. Every other tool (including SvABA’s filtered output) had a recall rate of less than 10% (**Fig. 4);** recall for Pindel could not be computed as it did not output interchromosomal SVs, which constituted the known SVs. At the coordinate level, recall rates for both SvABA’s unfiltered output and Delly drop to 17% and 13%, respectively. Thus, while SvABA’s unfiltered output can identify more known SVs at the chromosomal level, it performs worse at the coordinate level and is similar to Delly’s output.

For each cell line, **Supp Table 2** indicates which SV calling tool detected the cell line’s experimentally determined SV. 11 SVs were detected by both Delly and SvABA only (**Supp Table 2**). On one hand, there were 32 known SVs only called by SvABA (i.e., not Delly) (**Supp Table 2**) On the other hand, there were 2 SVs that were detected by Delly but not by SvABA. There were 10 known SVs not detected by any tool (**Supp Table 2**). Four known SVs were detected by at least three tools. No known SVs were detected by all five tools.

A cluster analysis of each cell line’s recall of known SVs **(Supp Fig. 2a**) showed 2 groups of cell lines. Cluster Group 1 had a mean recall rate of 14% across all tools, 73% for SvABA, 0% for Delly, and 36.5% across SvABA and Delly. Cluster Group 2 had a mean recall rate of 35% across all tools, 87.5% for SvABA, 94% for Delly, and 91% across SvABA and Delly. Clearly, the detector tools performed worse with identifying the known SVs in the first cluster group of cell lines. Another key takeaway is that none of Cluster Group 1’s known SVs were detected by Delly but 94% of Cluster Group 2’s known SVs were detected by Delly. Each cluster group’s cell lines are indicated in the supplementary materials.

A second cluster analyses (**Supp Fig. 2b**) on each tool’s recall resulted in three clusters, one with SvABA (78% recall at the chromosomal level, **Fig. 4**), another cluster containing Delly (28% recall at the chromosomal level, **Fig. 4**), and a third cluster defined by the remaining tools (BreakDancer, Smoove, Manta) who have a recall of less than 20% at the chromosomal level (**Fig. 4**).

### Comparison of SV tool overlap

Recall rates were computed for BreakDancer, Delly, Smoove, LUMPY, SvABA’s filtered and unfiltered output **(Fig. 4)**. Ultimately, we examined the amount of overlap in SV call among four tools: BreakDancer, Delly, SvABA’s unfiltered output and Smoove. LUMPY was not included because Smoove is a more advanced version of LUMPY and both tools had the same recall (3%, **Fig. 4**). SvABA’s filtered output was not included because SvABA’s unfiltered output had a much superior recall (2% vs 78%, **Fig. 4**). Pindel was not included because its recall could not be computed because it did not output interchromosomal SVs.

Shown in **Fig. 5** is the number of overlapping SVs for a given criterion. The first defined criterion was the number of tools that identified a given SV to quantify the amount of robustness (to tool) in SVs called. The number of overlapping SVs decreased by an order of magnitude when an additional tool was required (10,000s for 2 tools, 1,000s for 3 tools, 100s for 4 tools, 10s for 5 tools). The second defined criterion allowed base pair variation between two tools’ calls for a given SV to quantify positional variation. By applying this criterion, the number of overlapping SVs decreased by around a factor of 2 when the allowed base pair variation decreased by an order of magnitude. For example, for criteria involving 3 tools, an error range of 10bps yielded 2,198 SVs and the error range of 100bps yielded 4,375 SVs. **Supp Fig. 3** visually shows overlapping SVs for criterion that yielded less than 1000 SVs.

### Comparison of tools top SV genes

Except for Delly, each tool’s 10 most frequent genes had counts around 100 **(Fig. 6)**. The counts of Delly’s top 10 genes were an order of magnitude larger (i.e., thousands) **(Fig. 6a)**. There were 5 overlapping genes present in at least two tools’ 10 most frequent genes: TTC27, VPS13B, CTNNA, FGF1, and PDE11A. The gene TTC27 had 8695 and 115 occurrences in Delly and SvABA SV calls, respectively. It has been shown to be a topv50 up-regulated gene in the MM cell lines KMS11, MM.1S and OPM1^36^. The gene VPS13B had 102 and 76 occurrences in SvABA and Smoove’s SV calls, respectively. This gene^a^ has a low copy number in the MMM1S cell line relative to other cell lines from the CCLE Cell Line Gene CNV Profiles dataset. The genes CTNNA3, FGF14, and PDE11A all show a high frequency in SV calls among all the cell lines by both BreakDancer and Delly. The gene CTNNA3 has been shown to play a role in cell-to-cell adhesion in multiply myeloma^b^. FGF14 is an inclusion criterion in 1 clinical trial for multiple myeloma^c^. The gene PDE11A is involved in prostate^d^ and kidney cancer^e^.

### Concordance of tool SV genes

Upon first looking at (**Fig. 7)**, it appears that any tool subset containing BreakDancer had an especially low number of overlapping genes. For example, the gene overlap in BreakDancer, Smoove and SvABA (588 genes) was small compared to the gene overlap in Delly, Smoove and SvABA’s output (3201 genes).This can be attributed to the fact that the number of genes in BreakDancer’s output is 1 fewer order of magnitudes than the other tools. Thus, below we describe the overlap of non-BreakDancer tools separate from the overlap among all tools.

With respect to the overlap between Delly, Smoove and SvABA, two pairs yielded more than ten thousand genes: Delly and Smoove, Delly and SvABA. The other pair (Smoove and SvABA) yielded 3,301 overlapping genes. This was similar to the number of genes overlapping among these tools: 3,291.

With respect to BreakDancer, nearly all its identified genes were also present in Delly; only 7 genes detected in BreakDancer were not detected by SvABA. 70% of BreakDancer’s genes were also present in SvABA’s genes and a much smaller, 29% also overlapped with Smoove. Intriguingly, the gene overlap of the triplet BreakDancer, Delly and SvABA was only two less than between the pair BreakDancer and SvABA. All other tool combinations yielded less than 1000 overlapping genes. No genes were identified by all the tools.

## Discussion

Even though SV calling tools are designed and benchmarked on WGS data^23-25^, they are predominantly applied to cheaper WES data^1,6-22^. To our knowledge, this is the first benchmarking study of multiple SV calling tools on WES data. On 67 Multiple Myeloma (MM) cell lines, none of the seven evaluated tools (Delly, SvABA, BreakDancer, Smoove, LUMPY, Manta, Pindel) identified more than 20% of known SVs at the genomics coordinate level. This implies that any SVs identified by applying these SV caller tools to WES data should be confirmed experimentally before being declared legitimate. Due to the large heterogeneity present in the output of these seven SV callers, it is possible that SVs identified by multiple SV callers may be more likely to be experimentally confirmed than SVs identified by a single caller. As it is not clear whether these SV callers would recall known (i.e., experimentally determined) SVs in other cancers, future work should also evaluate these workflows on WES data from other cancers, ideally one cancer at a time.

## Conclusions

The poor recall of known SVs by multiple SV callers in this benchmarking study suggests one cannot merely treat the output of SV callers on WES data as immediately legitimate. Considering that WES data continues to drive many clinical grade sequencing panels for molecular tumor profiling to inform on targeted therapy and as yet, there is no unequivocal SV caller that has proven optimal for use with WES data on all cancer types, it is critical that we start to explore the performance of SV workflows applied to WES, one cancer at a time.

## Data Availability

Independent processed data is available at www.keatslab.org and raw data is available from Dr. Jonathan Keats upon request. All samples were de-identified. All code used to generate results can be found at https://github.com/kmlabdms/SV_MM/

https://www.keatslab.org/myeloma-cell-lines

https://github.com/kmlabdms/SV_MM/

## Declarations

### Ethics approval and consent to participate

This study used previously established human multiple myeloma cell lines obtained from Dr. Jonathan Keats lab (https://www.keatslab.org/myeloma-cell-lines) at the Translational Genomics Research Institute. No participants were recruited, and no new patient specimens were collected.

### Consent for publication

Not applicable. This manuscript reports cell line research and contains no identifiable individual patient information, images, or case descriptions.

## Availability of data and materials

Independent processed data is available at www.keatslab.org and raw data is available from Dr. Jonathan Keats upon request. All code used to generate results can be found at https://github.com/kmlabdms/SV_MM/

## Authors’ contributions

JKM conceived of the idea, designed the study, and drafted the initial manuscript.

DA directed computational methods and analyses.

LP led the analyses, generated figures, and helped to draft the manuscript

LP, RN, RH, RP, DN, DA performed the analyses. RN conducted literature review.

MY curated data and resources, performed reproducibility checks and edited manuscript.

## Acknowledgments

We thank Dr. Jonathan Keats from the Translational Genomics Research Institute for providing the raw FASTQ files from WES on the 67 Multiple Myeloma Cell lines. We also thank Jensen Richardson, who aligned the cell lines to the reference genome.

## Footnotes

a https://maayanlab.cloud/Harmonizome/gene_set/MM1S/CCLE+Cell+Line+Gene+CNV+Profiles

b https://www.ncbi.nlm.nih.gov/pmc/articles/PMC8

c https://www.mycancergenome.org/content/gene/fgf14/

d http://www.cancerindex.org/geneweb/PDE11A.htm

e https://www.proteinatlas.org/ENSG00000128655-PDE11A

## References

1 Mahmoud M, Gobet N, Cruz-Dávalos DI, Mounier N, Dessimoz C, Sedlazeck FJ. Structural variant calling: the long and the short of it. Genome biology. 2019 Nov 20;20(1):246.

2 Rausch T, Zichner T, Schlattl A, Stütz AM, Benes V, Korbel JO. DELLY: structural variant discovery by integrated paired-end and split-read analysis. Bioinformatics. 2012 Sep 15;28(18):i333–9.

3 Wala, J. A., Bandopadhayay, P., Greenwald, N. F., O’Rourke, R., Sharpe, T., Stewart, C., … & Beroukhim, R. (2018). SvABA: genome-wide detection of structural variants and indels by local assembly. Genome research, 28(4), 581–591.

4 Chen K, Wallis JW, McLellan MD, Larson DE, Kalicki JM, Pohl CS, McGrath SD, Wendl MC, Zhang Q, Locke DP, Shi X. BreakDancer: an algorithm for high-resolution mapping of genomic structural variation. Nature methods. 2009 Sep;6(9):677–81.

5 Layer RM, Chiang C, Quinlan AR, Hall IM. LUMPY: a probabilistic framework for structural variant discovery. Genome biology. 2014 Jun 26;15(6):R84.

6 Wang TY, Yang R. ScanITD: detecting internal tandem duplication with robust variant allele frequency estimation. Gigascience. 2020 Aug;9(8):giaa089.

7 Maroilley T, Tarailo-Graovac M. Uncovering missing heritability in rare diseases. Genes. 2019 Apr 4;10(4):275.

8 Manheimer KB, Patel N, Richter F, Gorham J, Tai AC, Homsy J, Boskovski MT, Parfenov M, Goldmuntz E, Chung WK, Brueckner M. Robust identification of deletions in exome and genome sequence data based on clustering of Mendelian errors. Human mutation. 2018 Jun;39(6):870–81.

9 Choi J, Manzano A, Dong W, Bellone S, Bonazzoli E, Zammataro L, Yao X, Deshpande A, Zaidi S, Guglielmi A, Gnutti B. Integrated mutational landscape analysis of uterine leiomyosarcomas. Proceedings of the National Academy of Sciences. 2021 Apr 13;118(15):e2025182118.

10 Roberts ND. Patterns of somatic genome rearrangement in human cancer (Doctoral dissertation).

11 Szymansky A, Kruetzfeldt LM, Heukamp LC, Hertwig F, Theissen J, Deubzer HE, Willing EM, Menon R, Fuchs S, Thole T, Schulte S. Neuroblastoma risk assessment and treatment stratification with hybrid capture-based panel sequencing. Journal of Personalized Medicine. 2021 Jul 22;11(8):691.

12 Sherman, R. M. (2021). Discovering novel human structural variation from diverse populations and disease patients: an exploration of what human genomics misses by relying on reference-based analyses, Johns Hopkins University.

13 Zhou X. Human Diploid Assembly and Variant Detection Methods Based on Linked-reads Technology. Stanford University; 2019.

14 Oh, C. (2020). The Tangent Copy-Number Inference Pipeline for Cancer Genome Analyses.

15 Chapuy B, Stewart C, Dunford AJ, Kim J, Kamburov A, Redd RA, Lawrence MS, Roemer MG, Li AJ, Ziepert M, Staiger AM. Molecular subtypes of diffuse large B cell lymphoma are associated with distinct pathogenic mechanisms and outcomes. Nature medicine. 2018 May;24(5):679–90.

16 Chapuy B, Stewart C, Dunford AJ, Kim J, Wienand K, Kamburov A, Griffin GK, Chen PH, Lako A, Redd RA, Cote CM. Genomic analyses of PMBL reveal new drivers and mechanisms of sensitivity to PD-1 blockade. Blood, The Journal of the American Society of Hematology. 2019 Dec 26;134(26):2369–82.

17 Wienand K, Chapuy B, Stewart C, Dunford AJ, Wu D, Kim J, Kamburov A, Wood TR, Cader FZ, Ducar MD, Thorner AR. Genomic analyses of flow-sorted Hodgkin Reed-Sternberg cells reveal complementary mechanisms of immune evasion. Blood advances. 2019 Dec 10;3(23):4065–80.

18 Kang L, Liu Y, Jin Y, Li M, Song J, Zhang Y, Zhang Y, Yang Y. Mutations of MACF1, encoding microtubule-actin crosslinking-factor 1, cause spectraplakinopathy. Frontiers in Neurology. 2020 Jan 15;10:1335.

19 Zehir A, Benayed R, Shah RH, Syed A, Middha S, Kim HR, Srinivasan P, Gao J, Chakravarty D, Devlin SM, Hellmann MD. Mutational landscape of metastatic cancer revealed from prospective clinical sequencing of 10,000 patients. Nature medicine. 2017 Jun;23(6):703–13.

20 Lan X, Bao H, Ge X, Cao J, Fan X, Zhang Q, Liu K, Zhang X, Tan Z, Zheng C, Wang A. Genomic landscape of metastatic papillary thyroid carcinoma and novel biomarkers for predicting distant metastasis. Cancer science. 2020 Jun;111(6):2163–73.

21 Im SW, Sung CO, Kim KS, Cho NH, Kim YM, Kwon GY, Moon KC, Choi SY, Lim JS, Choi YJ, Jung SJ. Genomic landscape of young-onset bladder cancer and its prognostic implications on adult bladder cancer. Cancers. 2020 Jan 28;12(2):307.

22 Lord EC, Poulter JA, Webster AR, Sergouniotis P, Khan KN, Benke PJ, Friedman L, Ali M, Inglehearn CF, Toomes C. Mutations in SLC38A8 and FOXD1 in patients with nystagmus and foveal hypoplasia. Investigative Ophthalmology & Visual Science. 2017 Jun 23;58(8):2786-.

23 Carss Carss KJ, Baranowska AA, Armisen J, Webb TR, Hamby SE, Premawardhana D, Al-Hussaini A, Wood A, Wang Q, Deevi SV, Vitsios D. Spontaneous coronary artery dissection: insights on rare genetic variation from genome sequencing. Circulation: Genomic and Precision Medicine. 2020 Dec;13(6):e003030.

24 Biswas, R., et al. (2016). “Genomic profiling of multiple sequentially acquired tumor metastatic sites from an “exceptional responder” lung adenocarcinoma patient reveals extensive genomic heterogeneity and novel somatic variants driving treatment response.” Molecular Case Studies 2(6): a001263.

25 Bademci G, Abad C, Incesulu A, Rad A, Alper O, Kolb SM, Cengiz FB, Diaz-Horta O, Silan F, Mihci E, Ocak E. MPZL2 is a novel gene associated with autosomal recessive nonsyndromic moderate hearing loss. Human genetics. 2018 Jul;137(6):479–86.

26 Chapman MA, Lawrence MS, Keats JJ, Cibulskis K, Sougnez C, Schinzel AC, Harview CL, Brunet JP, Ahmann GJ, Adli M, Anderson KC. Initial genome sequencing and analysis of multiple myeloma. Nature. 2011 Mar 24;471(7339):467–72.

27 Li H, Handsaker B, Wysoker A, Fennell T, Ruan J, Homer N, Marth G, Abecasis G, Durbin R, 1000 Genome Project Data Processing Subgroup. The sequence alignment/map format and SAMtools. bioinformatics. 2009 Aug 15;25(16):2078–9.

28 Li, H. (2013). “Aligning sequence reads, clone sequences and assembly contigs with BWA-MEM.” arXiv preprint arXiv:1303.3997.

29 McKenna A, Hanna M, Banks E, Sivachenko A, Cibulskis K, Kernytsky A, Garimella K, Altshuler D, Gabriel S, Daly M, DePristo MA. The Genome Analysis Toolkit: a MapReduce framework for analyzing next-generation DNA sequencing data. Genome research. 2010 Sep;20(9):1297–303.

30 Chen X, Schulz-Trieglaff O, Shaw R, Barnes B, Schlesinger F, Källberg M, Cox AJ, Kruglyak S, Saunders CT. Manta: rapid detection of structural variants and indels for germline and cancer sequencing applications. Bioinformatics. 2016 Apr 15;32(8):1220–2.

31 Ye K, Schulz MH, Long Q, Apweiler R, Ning Z. Pindel: a pattern growth approach to detect break points of large deletions and medium sized insertions from paired-end short reads. Bioinformatics. 2009 Nov 1;25(21):2865–71.

32 Cingolani P. Variant annotation and functional prediction: SnpEff. InVariant calling: methods and protocols 2012 eb 24 (pp. 289–314). New York, NY: Springer US.

33 Cui Y, Chen X, Luo H, Fan Z, Luo J, He S, Yue H, Zhang P, Chen R. BioCircos. js: an interactive Circos JavaScript library for biological data visualization on web applications. Bioinformatics. 2016 Jun 1;32(11):1740–2.

34 Lex A, Gehlenborg N, Strobelt H, Vuillemot R, Pfister H. UpSet: visualization of intersecting sets. IEEE transactions on visualization and computer graphics. 2014 Nov 6;20(12):1983–92.

