## Supplementary material for "Benchmarking Structural Variant Detection in Whole-Exome Sequencing of 67 Multiple Myeloma Cell Lines": Figure Legends

**Figure 1. Analytical workflow for structural variant benchmarking and cell line characterization**
Sequencing reads from n = 67 Multiple Myeloma cell lines are aligned to the reference genome to generate BAM files, followed by SV identification across multiple calling algorithms. Callers are evaluated for recall against experimentally validated SVs. Then, the cell lines are characterized with ≈4,500 SVs, and are annotated to pinpoint target genes, with spatial distributions visualized via Circos plots.

**Figure 2. Quantitative and qualitative comparison of structural variant (SV) detection outputs across calling tools**
Total SVs detected, SV lengths, and SV types identified across a panel of n = 67 Multiple Myeloma cell lines.
**A)** Vertical bar chart showing the total number of SVs detected per tool (y-axis scale is logarithmic). Smoove detected much fewer SVs (n=6520) compared to BreakDancer (n=456,481), Delly (n=371,770), SvABA (n=5,999,091). Note that the y-axis corresponding to number of SVs is logarithmic.
**B)** Boxplots showing the distribution of SV length by detector tool on a logarithmic scale. The x-axis indicates individual SV detector tools. All tools except Smoove identified hundreds of thousands of SVs; Smoove identified thousands of SVs.
**C)** Horizontal bar chart showing proportion of detected SV types per tool. SvABA output consisted exclusively of breakends (BNDs, 100%). Smoove output demonstrated high heterogeneity in SV types: BNDs (28%), Deletions (DEL, 41%), Duplications (DUP, 29%), and Inversions (INV, 2%). Delly and BreakDancer output consisted primarily of deletions (94% and 95%, respectively). BreakDancer uniquely identified 'Other' SV types, including inter-chromosomal (CTX) and intra-chromosomal (ITX) translocations.

**Figure 3**. **Runtime independent of sample read count for the fastest SV detector tools**
Tool runtimes (excluding preprocessing) plotted against sample read count across n = 67 Multiple Myeloma cell lines.
**A)** Runtime for high-speed SV detector tools (Delly, BreakDancer, Manta, and Smoove; y-axis scale: 0-800s). Manta and Smoove exhibited the fastest runtimes (<200s).
**B)** Runtime for slower SV detector tools (SvABA and Pindel; y-axis scale: 5,000 – 25,000s).

**Figure 4**. **SvABA achieved significantly higher recall of experimentally determined SV compared to Delly, BreakDancer, Smoove, LUMPY, Manta**
Percentage recall of experimentally determined SVs evaluated across n = 67 Multiple Myeloma cell lines.
**Blue bars:** Chromosomal-level recall, defined as identifying an experimentally determined interchromosomal SV sharing the target starting and ending chromosomes. SvABA (78%) and Delly (28%) were the only callers to achieve >20% recall.
**Orange bars:** Coordinate-level recall, defined as identifying an SV within the expected genomic boundary interval (e.g., t(4;14)(p16.3;q32.3) in cell line KMSP183 bounded by chr4:0–4,500,000 and chr14:100,900,001–107,043,718). SvABA's recall is reduced from 78% at the chromosomal level to 19.5% at the coordinate level.

**Figure 5. Overlap of SV identified by N tools ± k bps decreases by an order of magnitude for every increase in N**
Total number of overlapping SVs identified as a function of caller consensus (N tools, x-axis) and allowed base-pair window variance (± *k* bps, color key; logarithmic y-axis). Increasing the required tool consensus (N) decreased overlapping SVs by an order of magnitude. Restricting the allowed base-pair variance by an order of magnitude reduced the number of overlapping SVs by approximately two-fold.

**Figure 6. Five genes overlap among each tool's top 10 most frequent genes**
**A) Top 10 most frequent genes in Delly output**
**B) Top 10 most frequent genes in SvABA output**
**C) Top 10 most frequent genes in BreakDancer output**
**D) Top 10 most frequent genes in Smoove output**
Frequency of gene involvement in SV calls across n = 67 Multiple Myeloma cell lines, annotated via SnpEff, shown for (**A**) Delly, (**B**) SvABA, (**C**) BreakDancer, and (**D**) Smoove. Five genes (TTC27, VPS13B, CTNNA, FGF1, and PDE11A) were shared among the top 10 lists of at least two detector tools. Numerical labels above each bar denote absolute SV counts per gene.

**Figure 7. Intersection of affected genes across SV caller subsets**
Concordance plot displaying genes overlaps across combinations of four SV callers. Dot matrix combinations indicate tool subsets; filled dots denote included tools. Subsets containing BreakDancer showed lower gene overlaps due to a lower overall magnitude of detected genes. Excluding BreakDancer, caller pairs Delly–Smoove and Delly–SvABA each shared >10,000 genes, while Smoove–SvABA shared 3,301 genes. Three-way consensus among Delly, Smoove, and SvABA yielded 3,291 overlapping genes. No single gene was identified simultaneously by all four tools.

**Supplementary Figure 1. K-means cluster analysis of runtime for each tool and each cell line**
Principal component analysis (PCA) bidimensional projections of runtime vectors
**A)** Clustering of SV callers based on runtime vectors across all cell lines (K = 5 clusters). Cluster 1 comprises Manta and Smoove (the two leftmost points) whose runtimes were by the fastest (<200s). Cluster 2 consists of SvABA (the green most vertically high dot). Cluster 3 consists of Pindel (the rightmost dot). Cluster 4 consists of Delly (the orange left dot). Cluster 5 consists of BreakDancer (the left dark red dot).
**B)** Clustering of Multiple Myeloma cell lines based on runtime profiles across all callers (k = 4 clusters). Cell lines were excluded if output was not available for all callers.

**Supplementary Figure 2**. **Clear K-means clustering of each tool's recall results, less clear at cell line level**
PCA bidimensional projections of recall metrics across cell lines with clinically validated SVs.
**A)** Cell line clustering based on recall vectors across tools (K = 2 clusters).
**B)** Tool clustering based on recall vectors across cell lines (K = 3 clusters). Cluster 1: SvABA (78% recall); Cluster 2: Delly (28% recall); Cluster 3: BreakDancer, Smoove, and Manta (<20% recall)

**Supplementary Figure 3. Circos plots of SVs identified by N tools ± k bps**
Circos plots depicting inter- and intra-chromosomal translocations (chromosomes 1–22, X, Y) yielding less than 1000 SVs.
**A) 3 tools ±0 bps; B) 4 tools ±10 bps; C) 4 tools ± 0bps**
**D) 5 tools ± 1000bps; E) 5tools ±100 bps; F) 5 tools ±10bps**
Only the translocations occurring in chromosomes 1-22 and the X or Y chromosomes were depicted. As illustrated in the upper left most Circos plot, visualizing more than 1000 SVs would result in a circle completely colored in red. Connecting links indicate inter-chromosomal translocations anchored at median breakpoint positions; non-connecting sector arcs represent intra-chromosomal rearrangements (e.g., Panel D illustrates isolated intra-chromosomal events on chromosome 1).
