## Supplementary Table 2 for "Benchmarking Structural Variant Detection in Whole-Exome Sequencing of 67 Multiple Myeloma Cell Lines"

**Supplementary Table 2**. **Detection of experimentally validated structural variants across individual Multiple Myeloma cell lines by caller.**

A choropleth table indicating the recall of known, experimentally validated SVs by six structural variant calling algorithms across evaluated Multiple Myeloma cell lines. While the majority of evaluated cell lines harbor a single experimentally determined benchmark SV, three cell lines (KMS28PE, MM1S, and JJN3) each contain two. Performance across the dataset demonstrated substantial caller variability: no single validated SV was detected simultaneously by all six tools, and only four known SVs were recovered by three or more tools. SvABA uniquely identified 32 validated SVs, Delly uniquely identified two, and 11 SVs were concordantly detected by SvABA and Delly alone. Conversely, ten experimentally validated SVs failed to be detected by any calling algorithm.

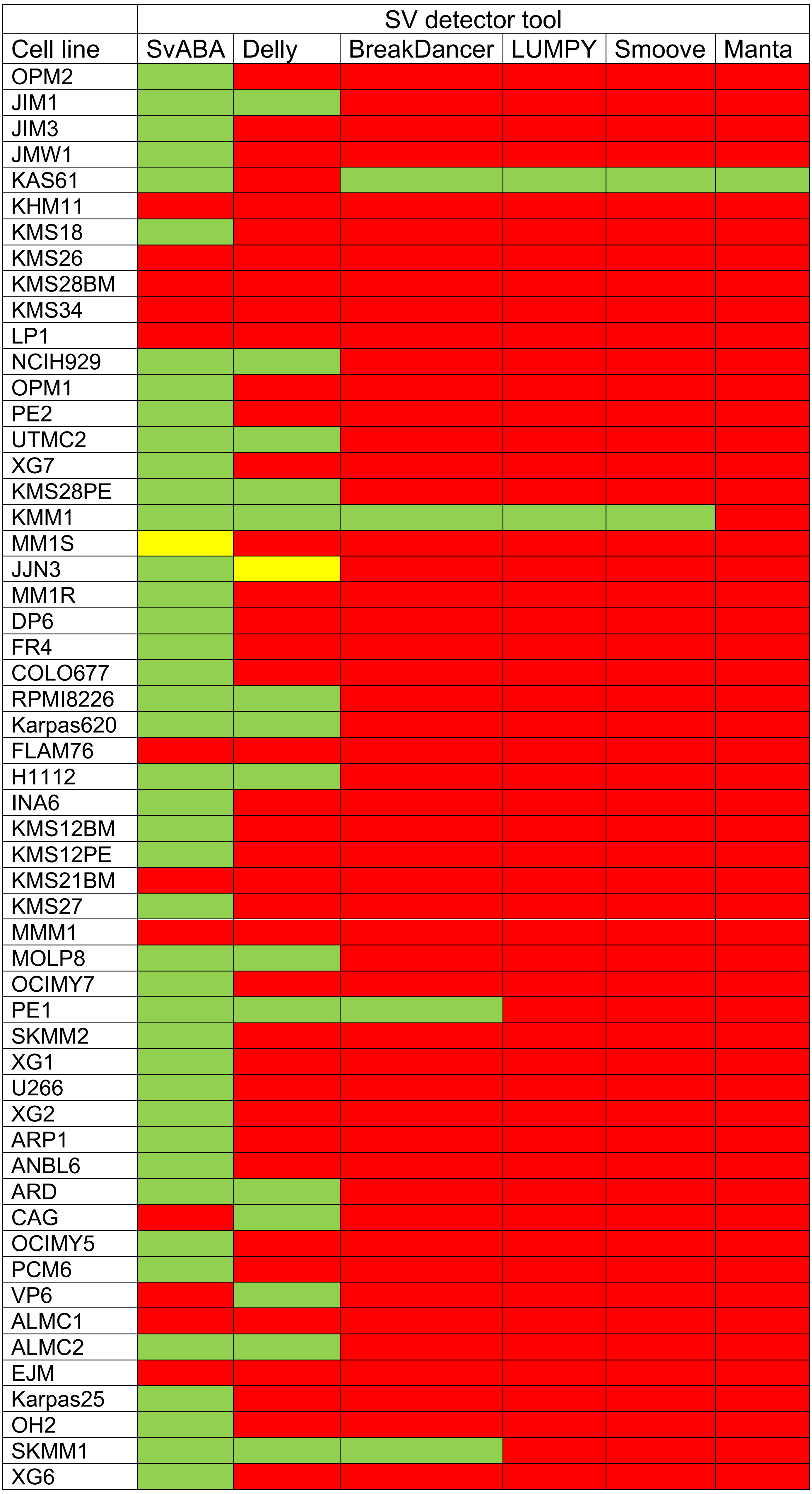
