## Supplementary material for "Benchmarking Structural Variant Detection in Whole-Exome Sequencing of 67 Multiple Myeloma Cell Lines": Figures

### Slide 1
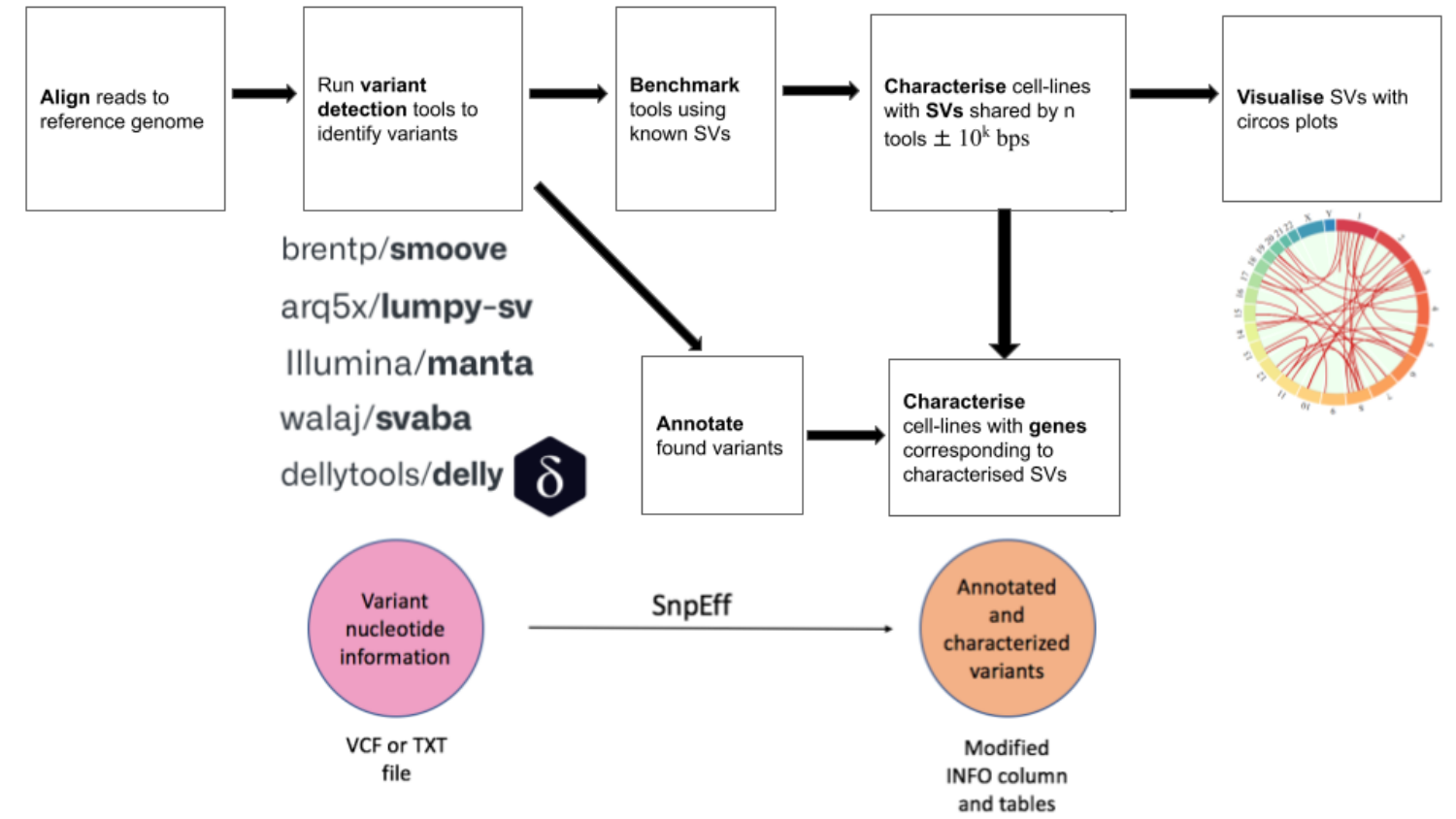

# ww

### Slide 2
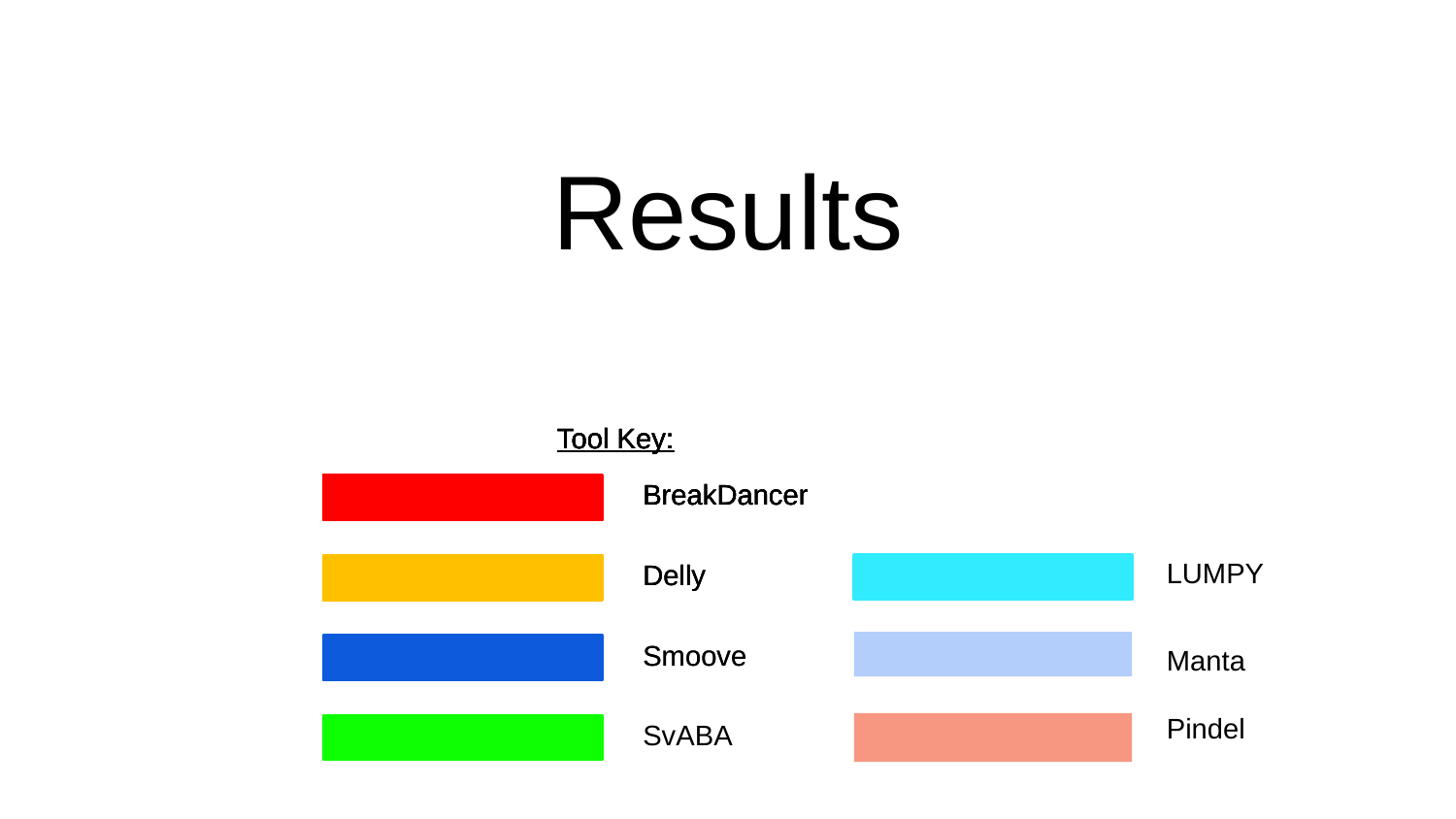

#### Results
Tool Key:
Tool Key:
Tool Key:
Tool Key:
Tool Key:
BreakDancer
BreakDancer
BreakDancer
BreakDancer
LUMPY
Delly
Delly
Delly
Smoove
Smoove
Manta
Pindel
SvABA

### Slide 3
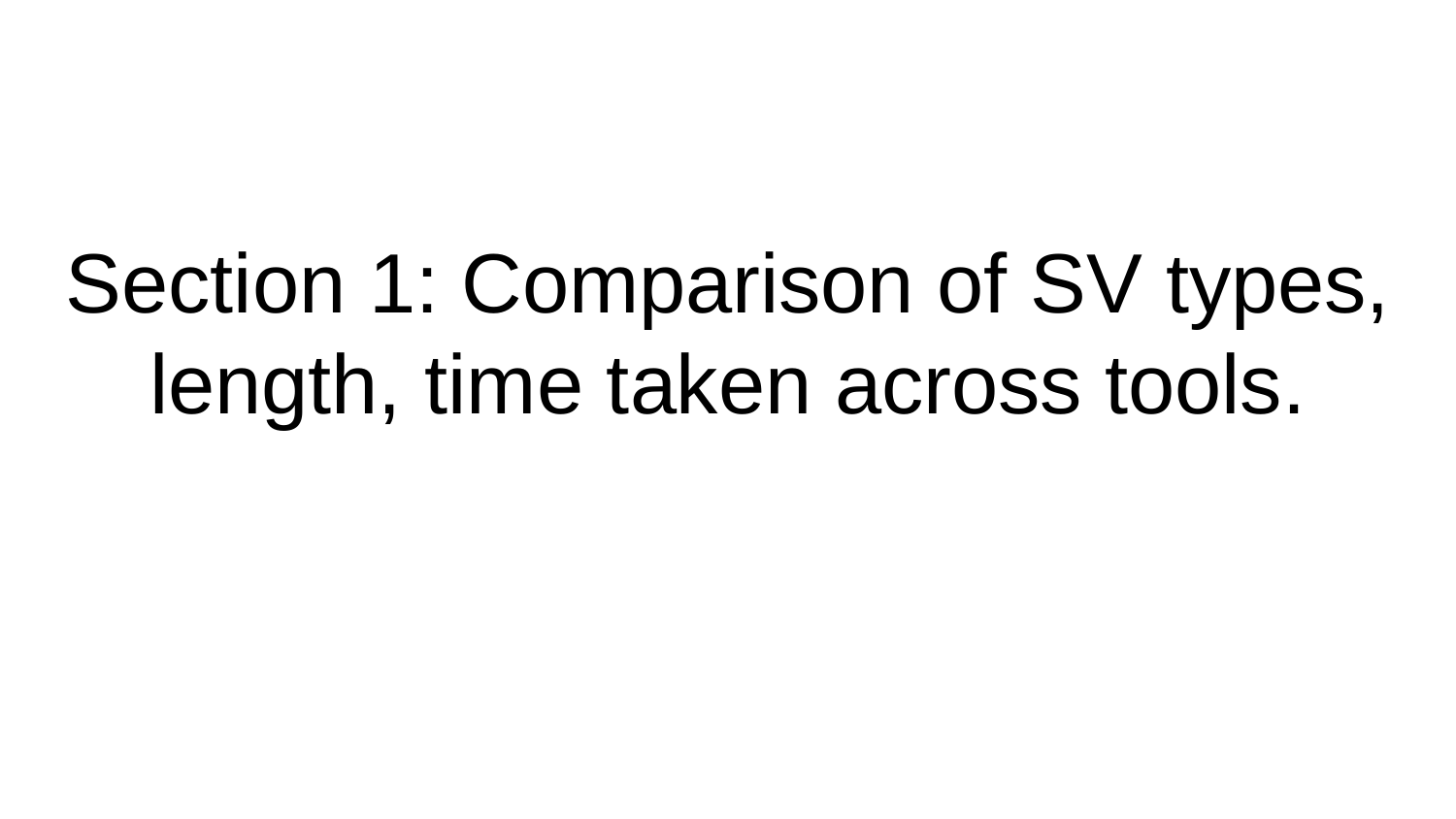

#### Section 1: Comparison of SV types, length, time taken across tools.

### Slide 4
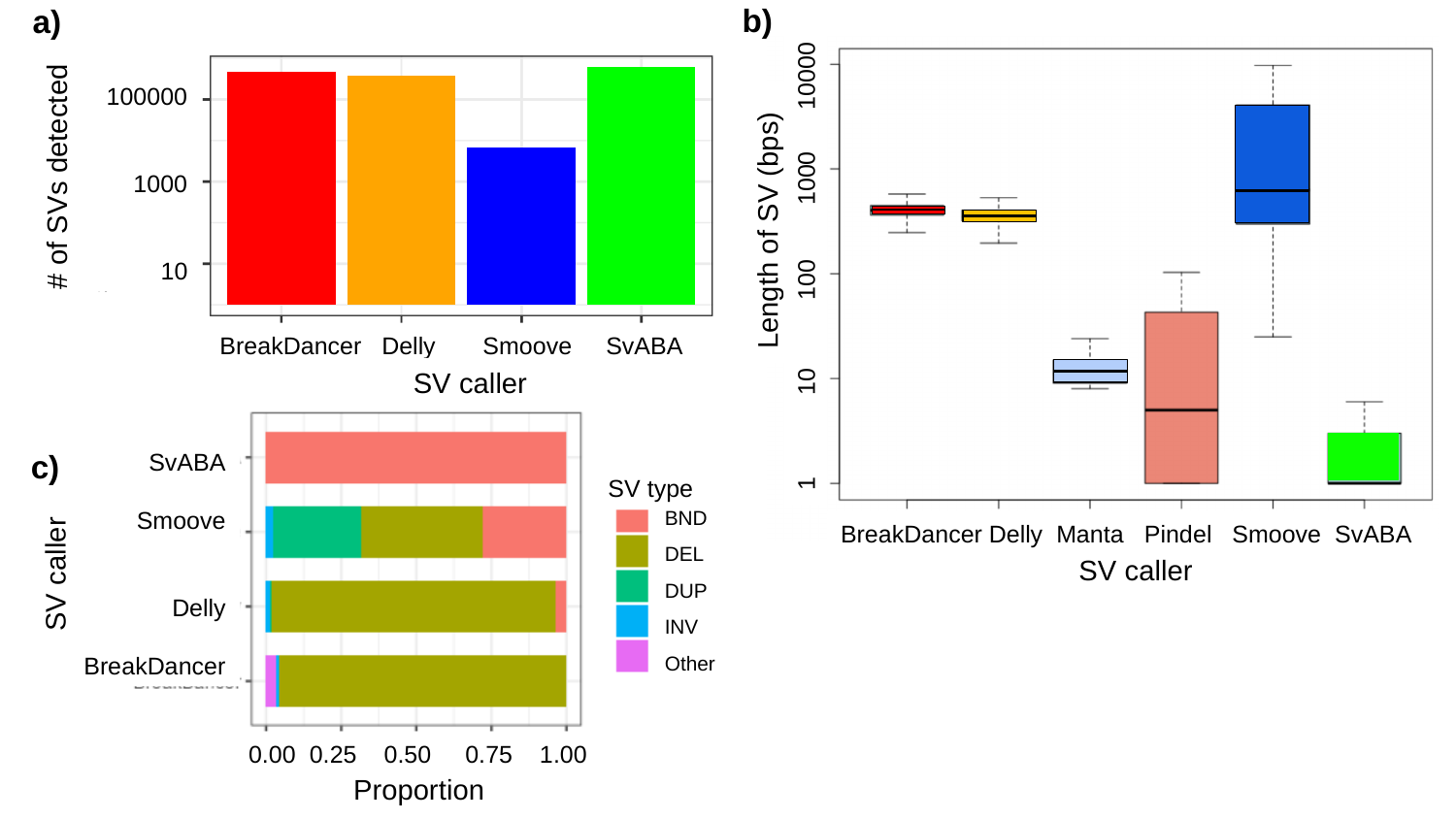

b)
a)
100000
1000
10
#### of SVs detected
(bps)
Length of SV (bps)
1 10 100 1000 10000
BreakDancer Delly Smoove SvABA
SV caller
c)
SvABA
Smoove
Delly
BreakDancer
SV type
BND
DEL
DUP
INV
Other
BreakDancer Delly Manta Pindel Smoove SvABA
SV caller
SV caller
0.00 0.25 0.50 0.75 1.00
Proportion

### Slide 5
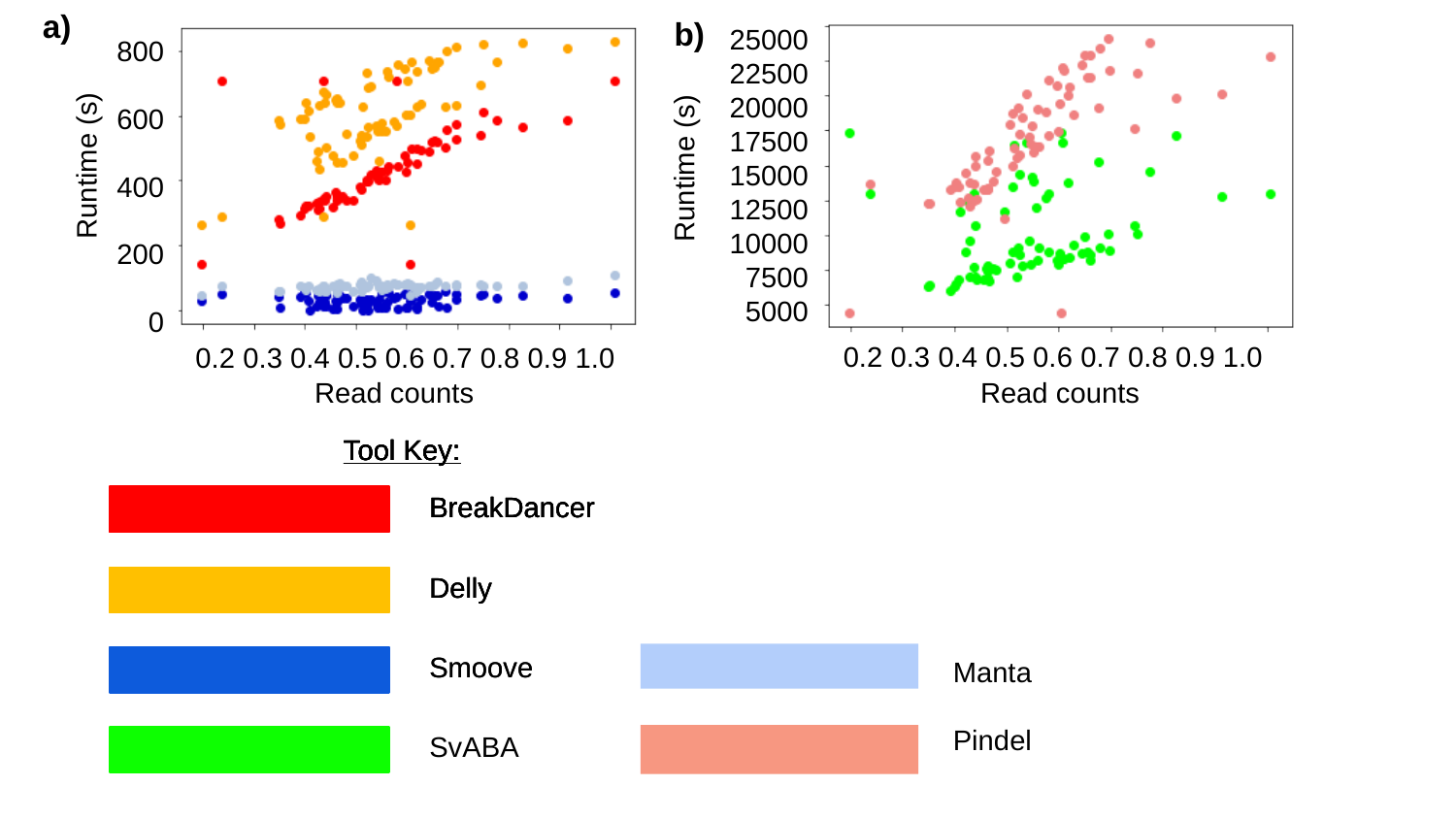

a)
b)
25000
22500
20000
17500
15000
12500
10000
7500
5000
b)
800
600
400
200
0
Runtime (s)
Runtime (s)
Runtime (s)
0.2 0.3 0.4 0.5 0.6 0.7 0.8 0.9 1.0
0.2 0.3 0.4 0.5 0.6 0.7 0.8 0.9 1.0
Read counts
Read counts
Tool Key:
Tool Key:
Tool Key:
Tool Key:
Tool Key:
BreakDancer
BreakDancer
BreakDancer
BreakDancer
Delly
Delly
Delly
Smoove
Smoove
Manta
Pindel
SvABA

### Slide 6
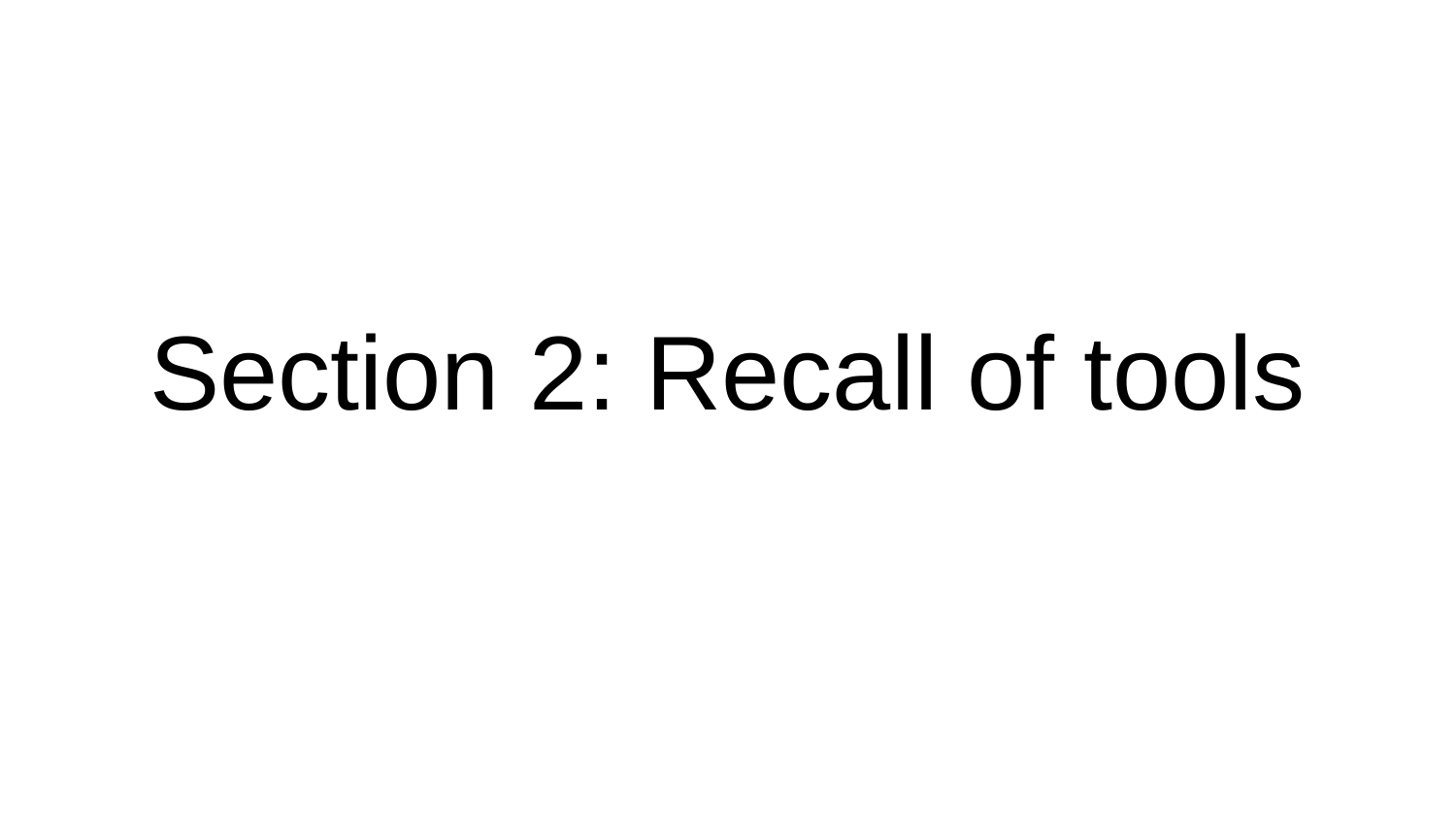

#### Section 2: Recall of tools

### Slide 7
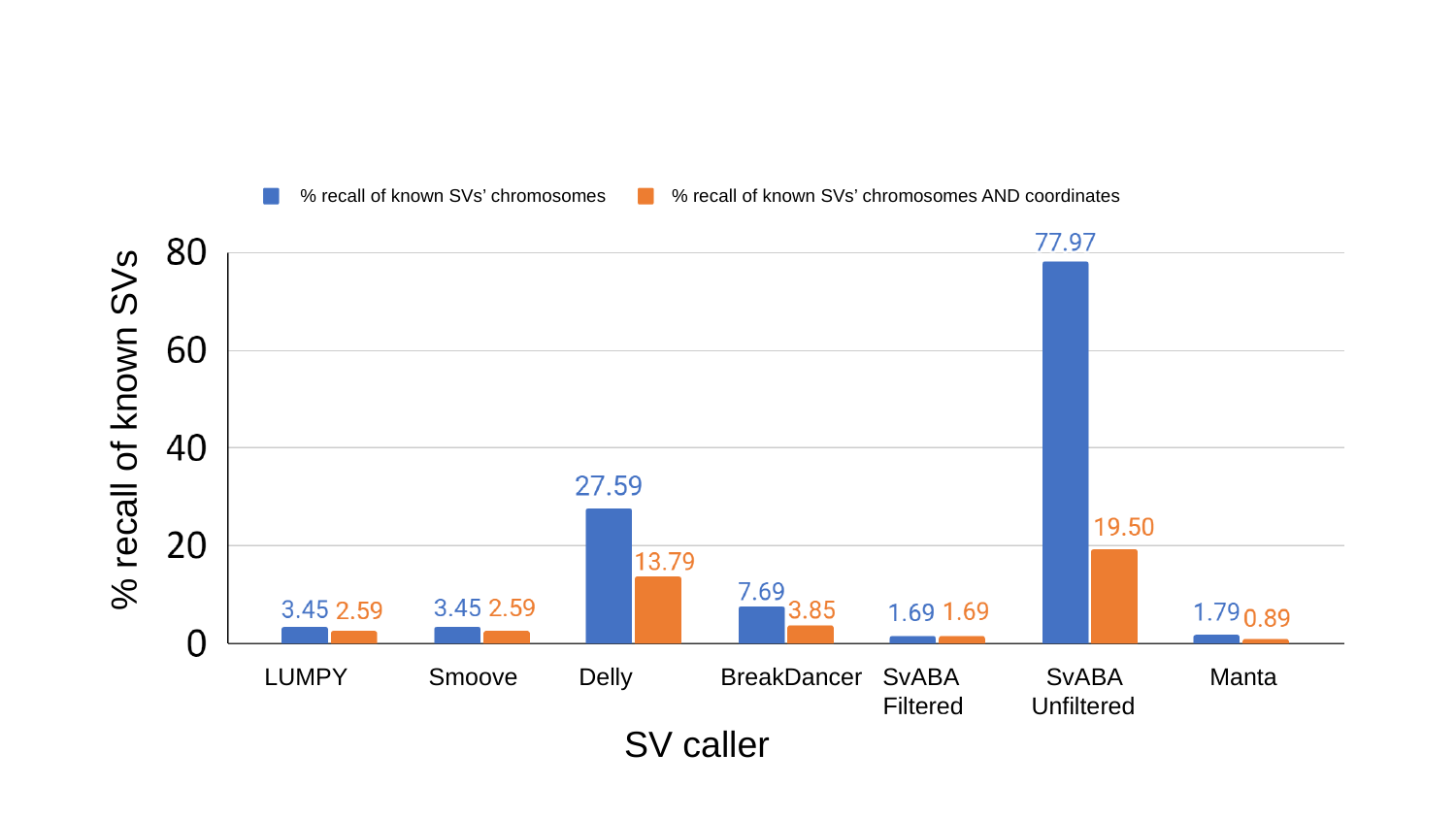

% recall of known SVs’ chromosomes
% recall of known SVs’ chromosomes AND coordinates
% recall of known SVs
LUMPY Smoove Delly BreakDancer SvABA SvABA Manta
 Filtered Unfiltered
SV caller
b)

### Slide 8
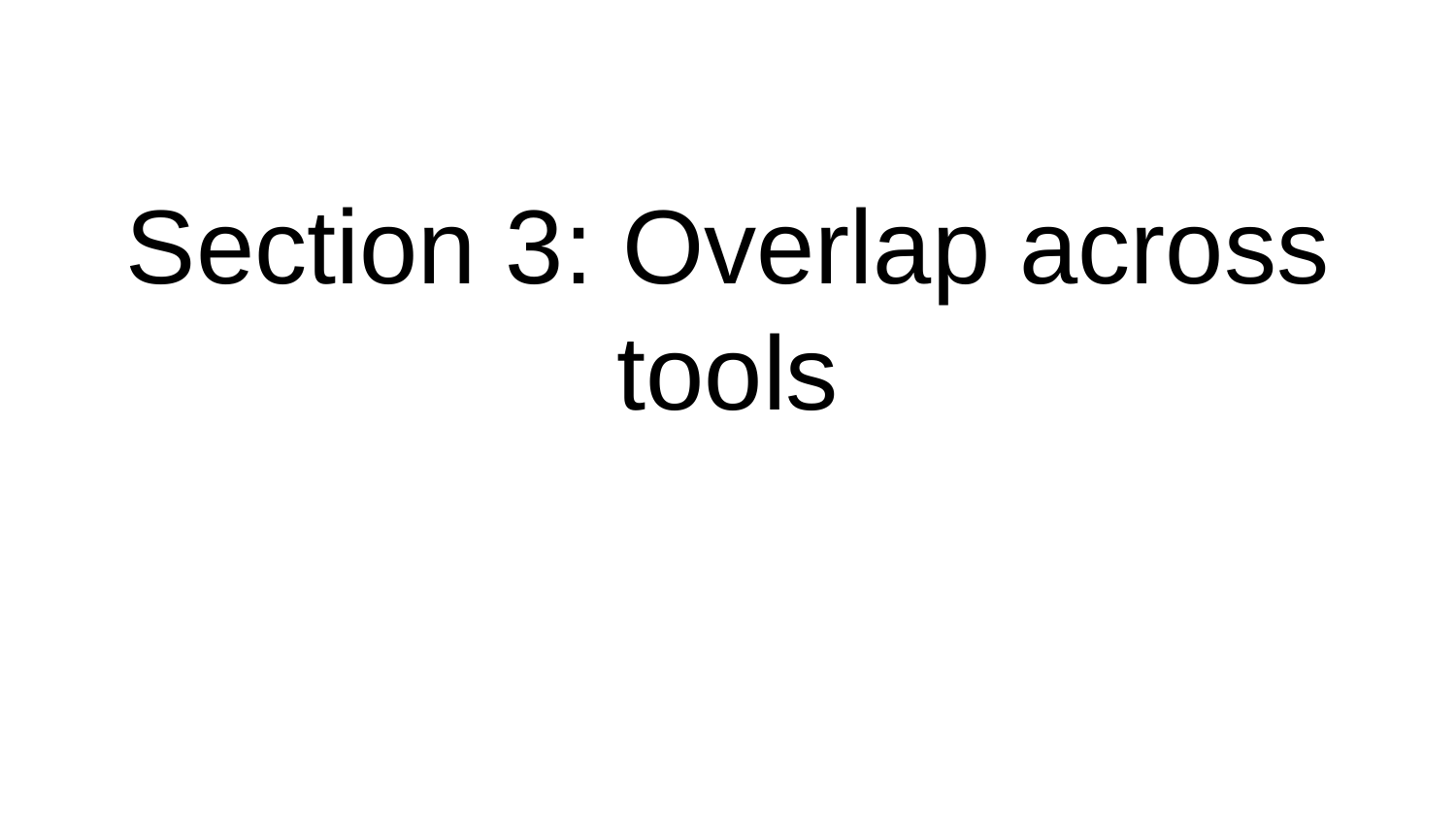

#### Section 3: Overlap across tools

### Slide 9
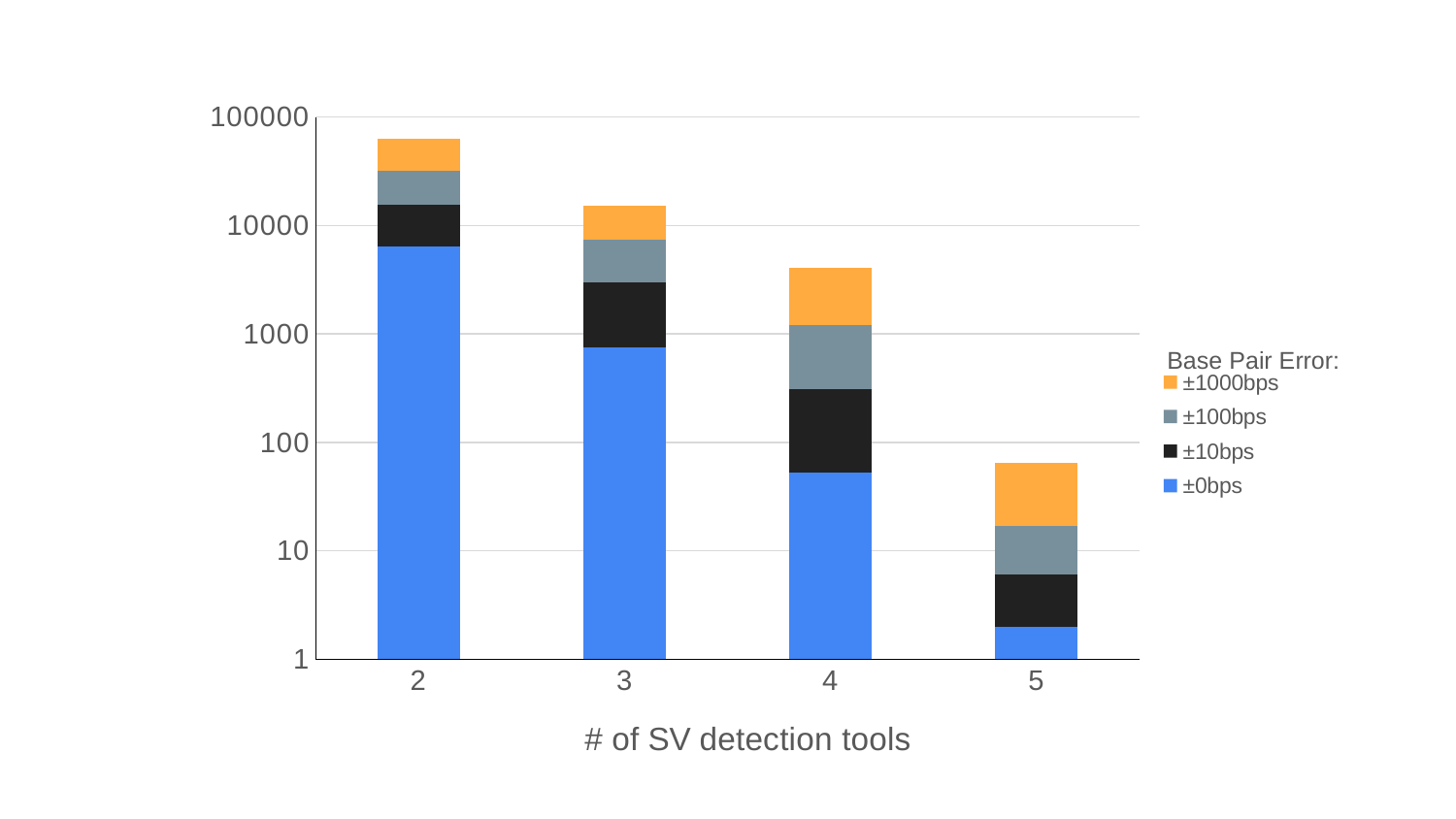

###### Chart
| Category | | | | |
|---|---|---|---|---|
| 2 | 6426.0 | 9042.0 | 16176.0 | 31928.0 |
| 3 | 758.0 | 2198.0 | 4375.0 | 7752.0 |
| 4 | 53.0 | 254.0 | 903.0 | 2868.0 |
| 5 | 2.0 | 4.0 | 11.0 | 48.0 |Base Pair Error:

### Slide 10
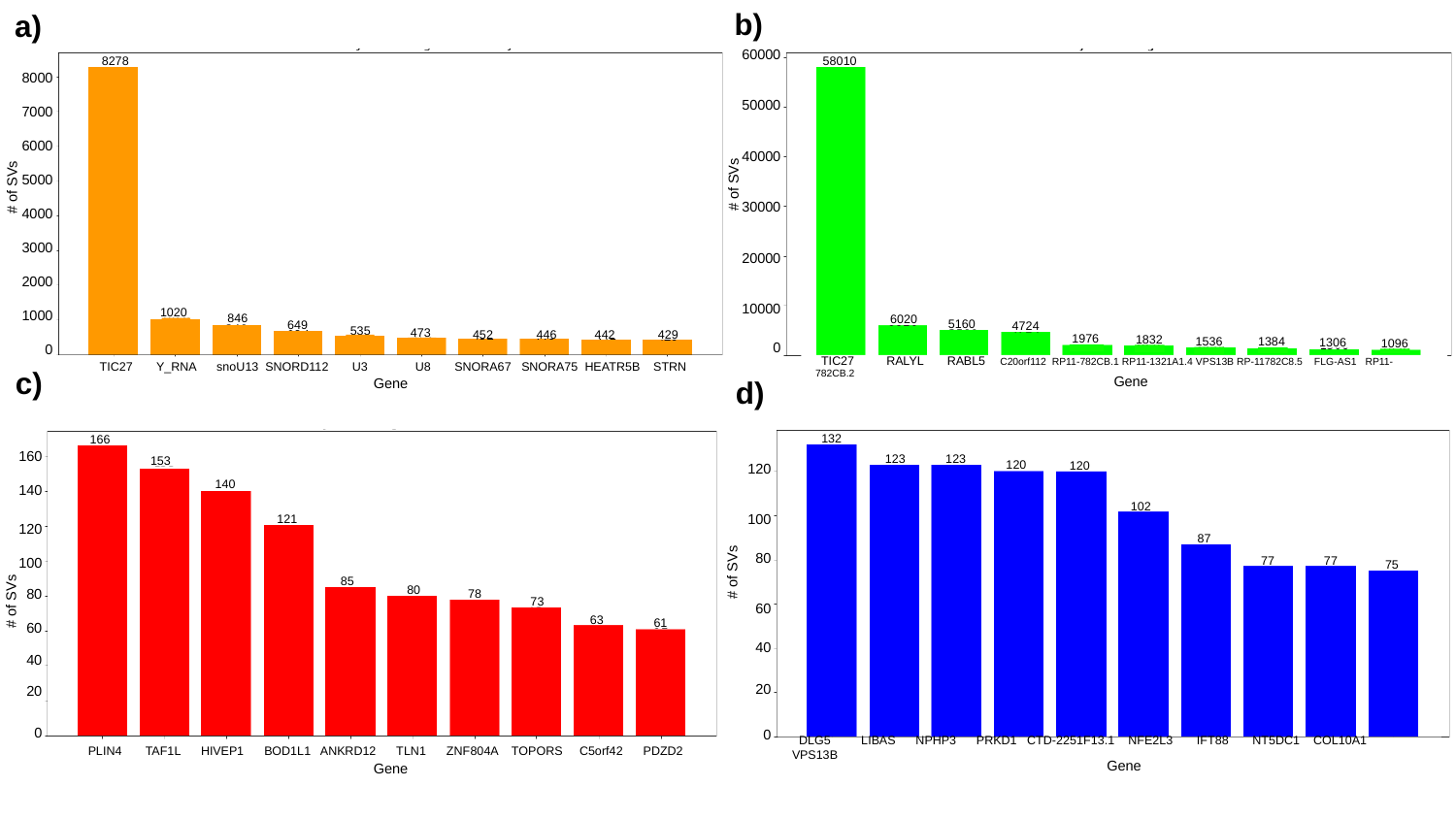

b)
a)
60000
50000
40000
30000
20000
10000
0
8278
58010
8000
7000
6000
5000
4000
3000
2000
1000
0
#### of SVs
#### of SVs
1020
846
649
6020
5160
4724
535
473
452
446
442
429
1976
1832
1536
1384
1306
1096
c)
 TIC27 RALYL RABL5 C20orf112 RP11-782CB.1 RP11-1321A1.4 VPS13B RP-11782C8.5 FLG-AS1 RP11-782CB.2
 TIC27 Y_RNA snoU13 SNORD112 U3 U8 SNORA67 SNORA75 HEATR5B STRN
d)
Gene
Gene
132
166
160
140
120
100
80
60
40
20
0
120
100
80
60
40
20
0
123
123
153
120
120
140
102
121
87
#### of SVs
77
77
75
85
#### of SVs
80
78
73
63
61
 DLG5 LIBAS NPHP3 PRKD1 CTD-2251F13.1 NFE2L3 IFT88 NT5DC1 COL10A1 VPS13B
 PLIN4 TAF1L HIVEP1 BOD1L1 ANKRD12 TLN1 ZNF804A TOPORS C5orf42 PDZD2
Gene
Gene

### Slide 11
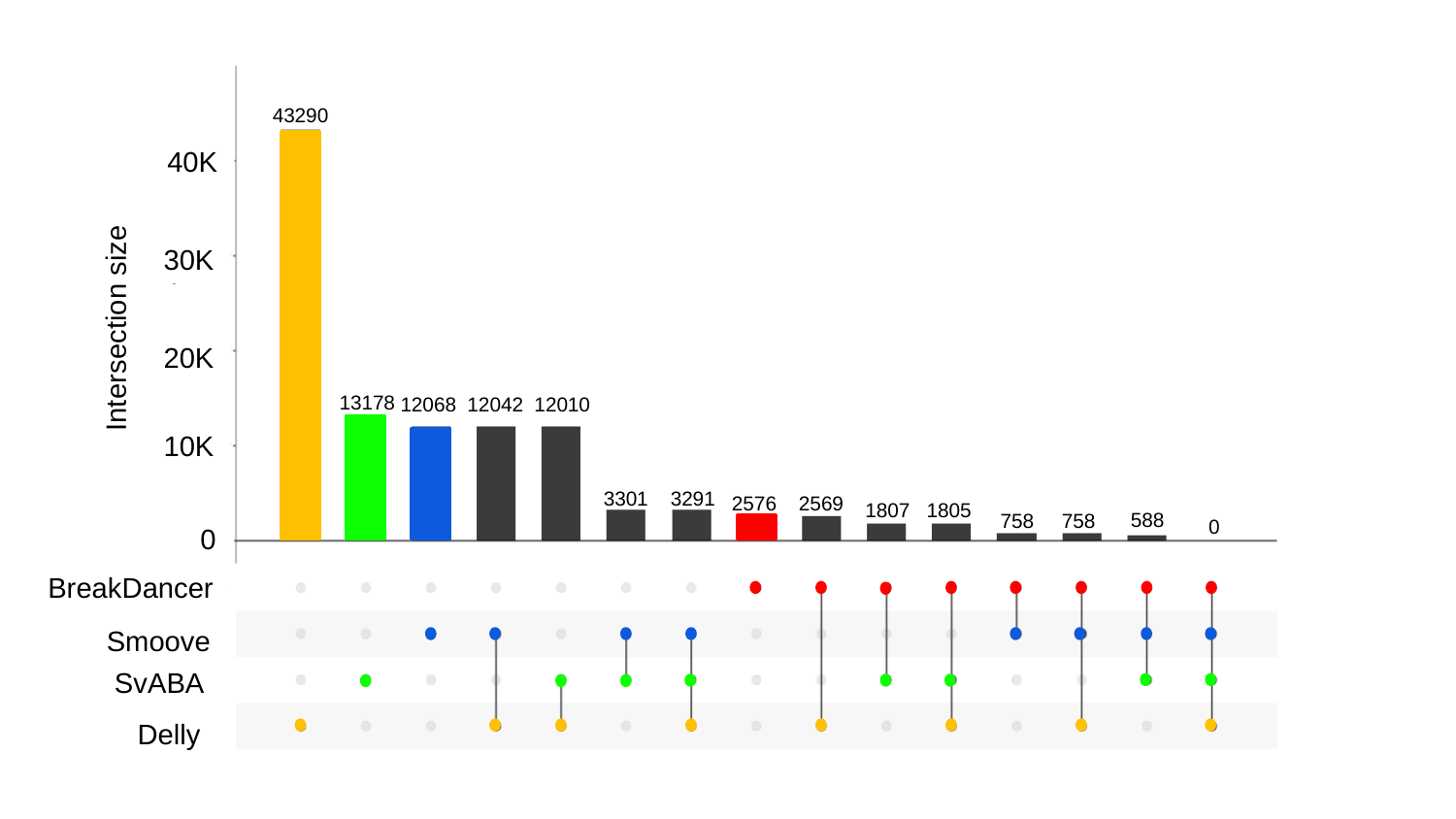

43290
40K
30K
Intersection size
20K
13178
12068 12042 12010
10K
2576 2569
3301 3291
1807 1805
0
588
758 758
0
BreakDancer
Smoove
SvABA
Delly

### Slide 12
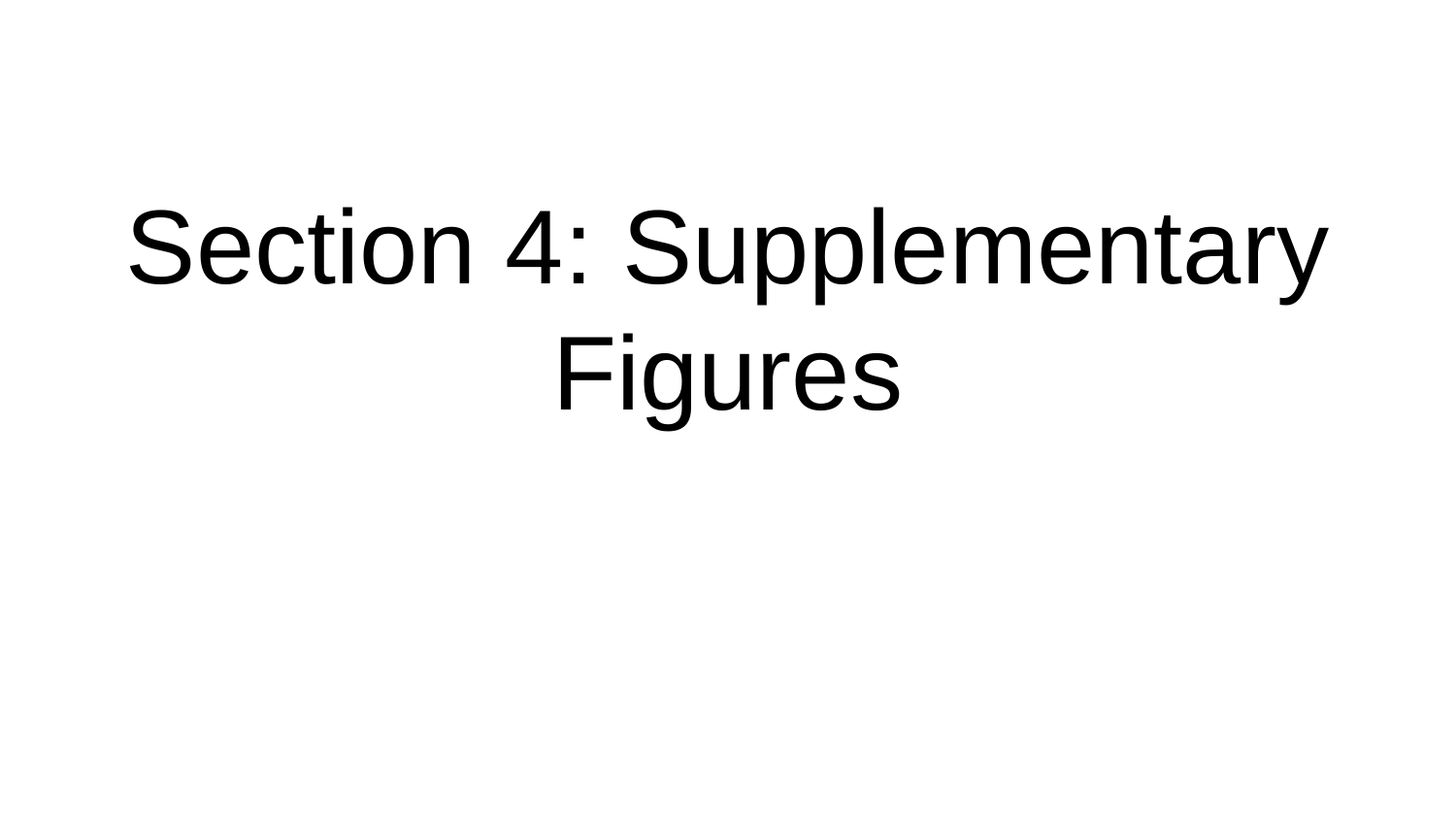

#### Section 4: Supplementary Figures

### Slide 13
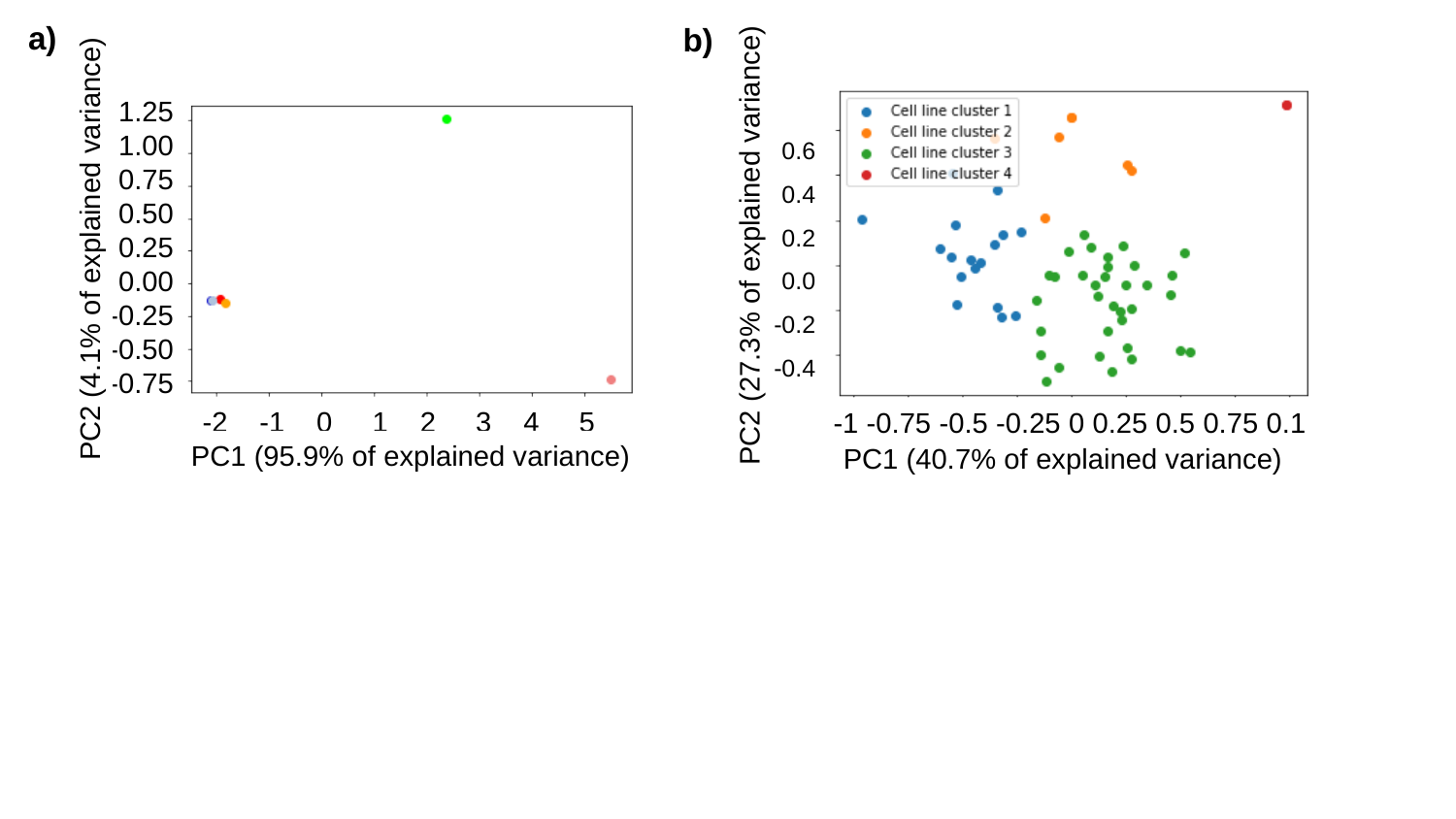

a)
b)
1.25
1.00
0.75
0.50
0.25
0.00
-0.25
-0.50
-0.75
0.6
0.4
0.2
0.0
-0.2
-0.4
PC2 (4.1% of explained variance)
PC2 (27.3% of explained variance)
-2 -1 0 1 2 3 4 5
-1 -0.75 -0.5 -0.25 0 0.25 0.5 0.75 0.1
PC1 (95.9% of explained variance)
PC1 (40.7% of explained variance)

### Slide 14
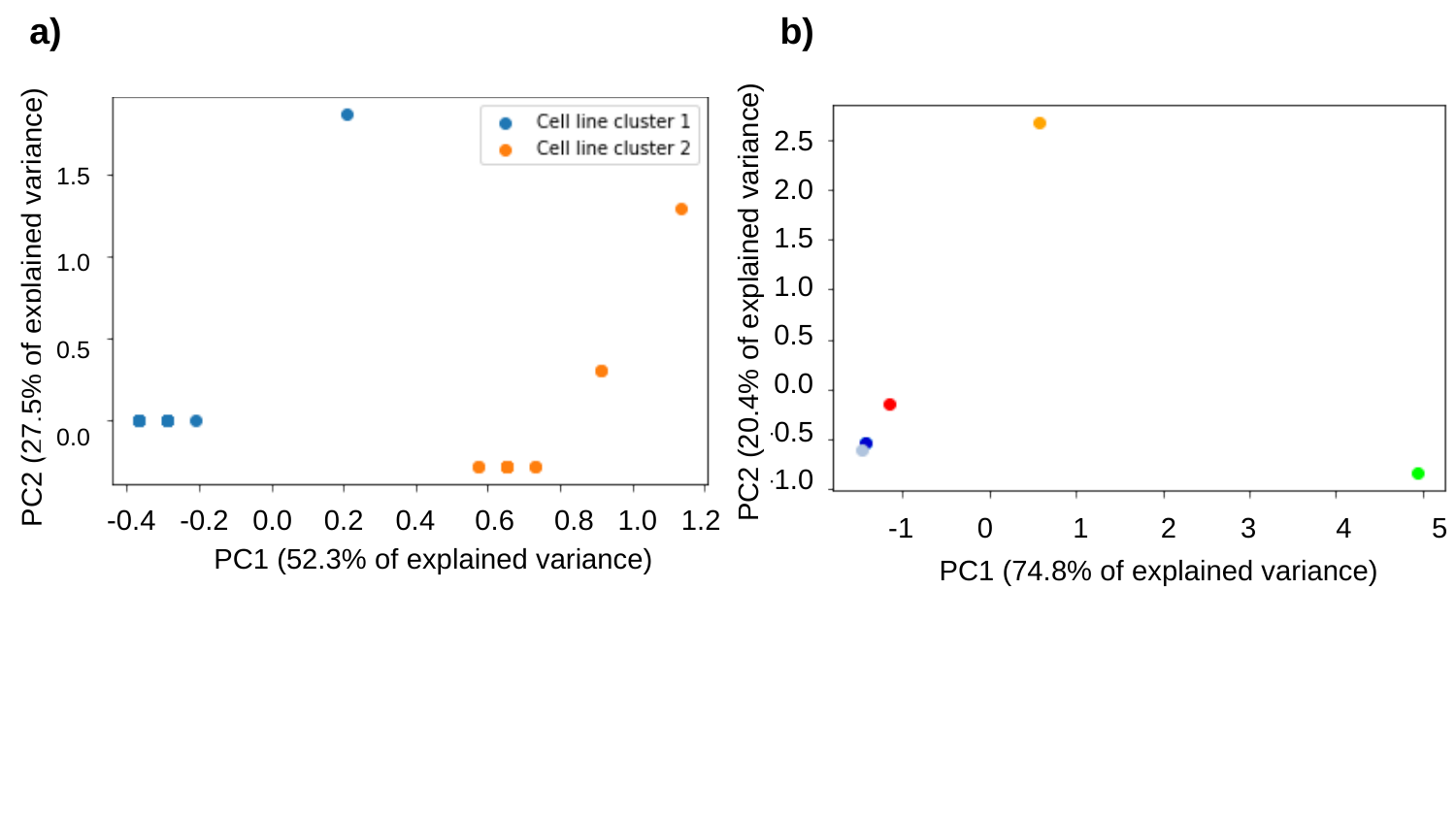

a)
b)
2.5
2.0
1.5
1.0
0.5
0.0
-0.5
-1.0
1.5
1.0
0.5
0.0
PC2 (20.4% of explained variance)
PC2 (27.5% of explained variance)
-0.4 -0.2 0.0 0.2 0.4 0.6 0.8 1.0 1.2
-1 0 1 2 3 4 5
PC1 (52.3% of explained variance)
PC1 (74.8% of explained variance)
-2 -1 0 1 2 3 4 5
-1 -0.75 -0.5 -0.25 0 0.25 0.5 0.75 0.1
PC1 (95.9% of explained variance)
PC1 (40.7% of explained variance)

### Slide 15
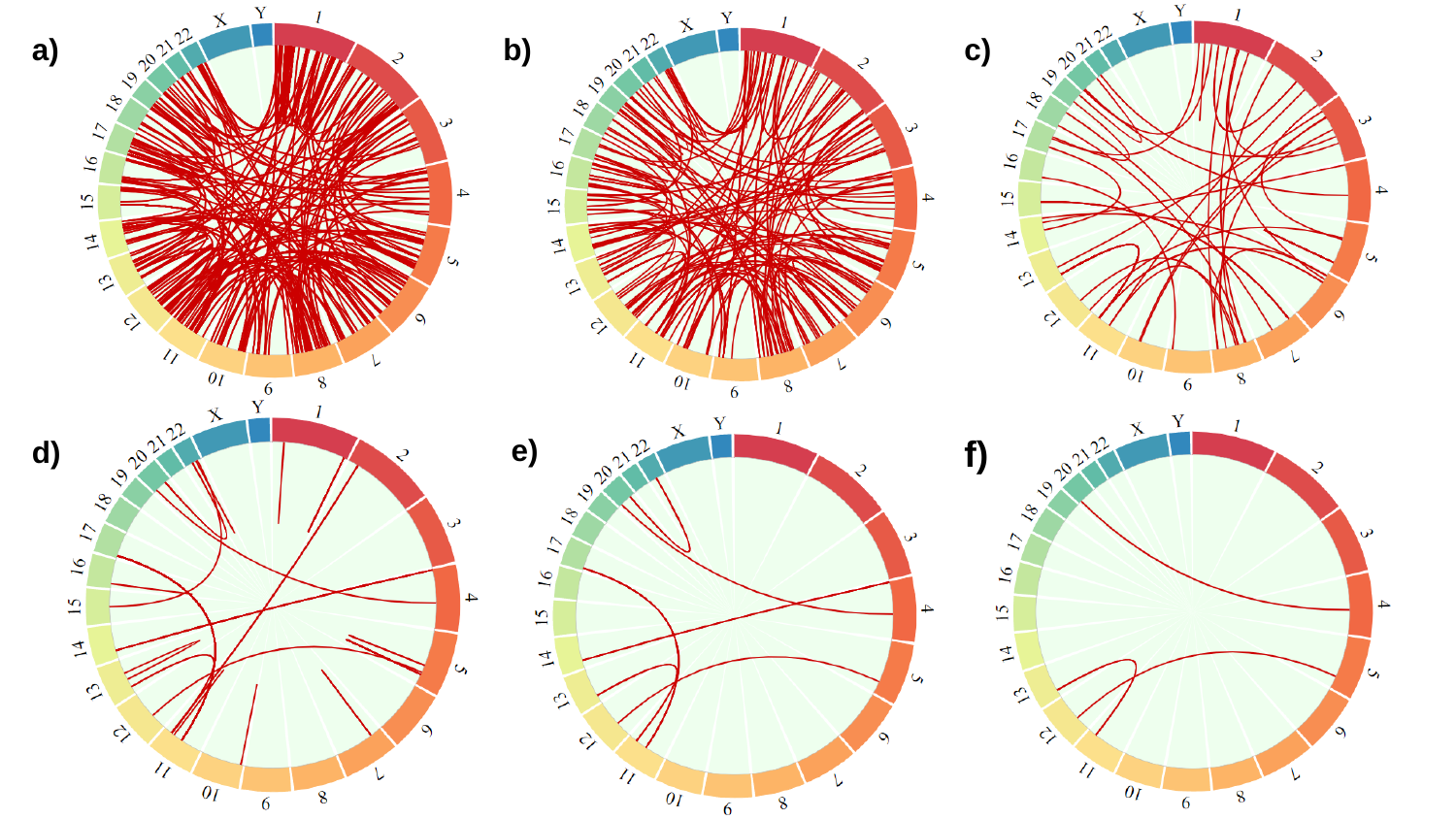

a)
b)
c)
f)
e)
d)
